# GLYCEMIC CONTROL AND PERIPHERAL ARTERIAL DISEASE IN DIABETIC FOOT PATIENTS

**DOI:** 10.1101/2024.04.01.24305179

**Authors:** María Patricia Aragón Carreño, Alfredo Aldama Figueroa, Fermín Rafael Martínez de Jesús, Aleksandar Cvetkovic-Vega, Jorge L. Maguiña Quispe

**Affiliations:** Escuela de Posgrado, Universidad Peruana Cayetano Heredia Lima, Perú; Unidad de Pie Diabético, Hospital Nacional Guillermo Almenara Irigoyen ESSALUD Lima, Perú; Instituto Nacional de Angiología y Cirugía Vascular, La Habana, Cuba; Centro de Prevención y Salvamento del Pie Diabético San Elián Veracruz, México; Escuela Profesional de Medicina Humana, Universidad Privada Antenor Orrego; Trujillo, Perú; Facultad de Ciencias de la Salud, Posgrado de Medicina, Universidad Científica del Sur, Lima, Perú

**Keywords:** ( DeCS): Glycemic control, Glycosylated Hemoglobin, Peripheral Arterial Disease, Diabetic foot, ankle-brachial index

## Abstract

**Introduction:** Diabetic foot is a severe complication of diabetes mellitus that can cause many amputations.

**Objectives:** To evaluate the association between glycemic control and peripheral arterial disease in patients with diabetic foot.

**Materials and Methods:** An analytical cross-sectional study was performed on patients diagnosed with diabetic foot with more than ten years of evolution of diabetes mellitus who were attended in the diabetic foot outpatient clinic of the Guillermo Almenara National Hospital between June 2015 and February 2017. Glycosylated hemoglobin (HbA1C) and ankle brachial index (ABI) were used to diagnose glycemic control and peripheral arterial disease, respectively. Generalized poisson log linear regression model was used to calculate the crude (cPR) and adjusted (aPR) prevalence ratios, with 95% confidence intervals, and a significance level of p < 0.05.

**Results:** 226 patients were included in the study, finding 23% (n=52) with good glycemic control, and 44.7% (n=101) with peripheral arterial disease. The adjusted model found statistically significant associations for glycemic control, age, and sedentary lifestyle with peripheral arterial disease. Compared with patients with good glycemic control, those with poor glycemic control had a 34% higher probability to present peripheral arterial disease (aPR: 1.34, 95%CI: 1.01-1.79, p=0.045).

**Conclusions:** The prevalence of good glycemic control in the population with diabetic foot is low. Poor glycemic control was found to be independently associated with the presence of peripheral arterial disease.

## INTRODUCTION

Diabetic foot (DF) is a severe complication of diabetes mellitus (DM) that can cause many amputations with a negative impact on the patient’s quality of life and high costs for the healthcare system. Worldwide, in 2016, the estimated prevalence of DF was 6.3%[1] and in 2017, in Latin America, 14.8% for nine countries in our region[2]. In Peru, between 2012 and 2020, there was reported an increasing trend from 5.9%[3], 10.8%[2] to 18.9%[4].

Peripheral arterial disease (PAD) is an occlusive atherosclerotic disease of the lower extremities, and its finding is a poor prognostic factor in patients with DF since it can lead to amputation of the affected extremity[5]. The onset and progression of PAD is related to inadequate glycemic control: for every 1% increase in glycosylated hemoglobin there is a 25% to 28% increase in the relative risk of PAD[6].

Although different studies have identified the benefits of strict glycemic control in diseases with microcirculatory damage such as retinopathy, nephropathy, and neuropathy, this situation has not been shown for macrocirculatory damage, such as the case of PAD in DF[7], despite the fact that the development of complications of this type increases cardiovascular morbidity and general mortality, as well as high economic and social costs[8].

The aim of this study is to evaluate the association between glycemic control and PAD in patients diagnosed with DF.

## MATERIALS AND METHODS

An analytical cross-sectional study was performed considering the population of all patients with DF and foot at risk who were attended in the DF outpatient clinic of the Guillermo Almenara Irigoyen National Hospital (HNGAI) between June 2015 and February 2017. As selection criteria, patients older than 18 years were included, with a time of type 2 DM greater than ten years of evolution and who agreed to participate in the study. Patients with type 1 DM, lupus, rheumatoid arthritis, vasculitis, Hepatitis B, C or HIV infections, stage V chronic kidney disease, recipients of some transplant, hospitalization in the last four weeks of evaluation, previous revascularization and amputees at the supracondylar or infracondylar level were excluded.

In addition, a non-probabilistic sampling was calculated, with a calculated statistical power of 94.59%, including 15% of losses and rejections, achieving a sample made up of 226 participants, considering the patient with a diagnosis of DF as the sampling unit, of which the most affected limb was selected.

The independent variable in this study was glycemic control, determined using the glycosylated hemoglobin (HbA1C) levels recorded in patients’ clinical histories at the time of their interviews. This was categorized based on the 2019[9] American Diabetes Association (ADA) criteria as Good Glycemic Control (HbA1C < 7%) or Poor Glycemic Control (HbA1C ≥ 7%).

The dependent variable was peripheral arterial disease (PAD) evaluated using the Ankle Brachial Index (ABI) his assessment adhered to the non-invasive vascular evaluation criteria set by the International Diabetes Federation’s Clinical Practice Recommendations of 2017[10,11]. An ABI value of ≤ 0.9 indicated the presence of PAD[12,13].

The ABI measurements were conducted using the NICOLET VERSALAB L.E Continuous Linear Doppler equipment, manufactured by VIASYS Healthcare in the USA in 2005. This equipment utilized an 8 MHz transducer, transmitter gel, and a blood pressure cuff.

Additionally, the study considered the following covariates : age, gender, current job, body mass index, time of evolution of type 2 DM, arterial hypertension, dyslipidemia, obesity, smoking, and sedentary lifestyle.

The study was conducted in two stages in the HNGAI DF Outpatient Clinic settings for 24 months with a frequency of 5 times per week from June 17^th^, 2015 to July 20^th^ 2017. In the first stage, each intervention included the obtaining of the written informed consent and completing the clinical history, within which palpation of foot pulses and ABI were considered, using the Ischemia Severity Scale (ISS)[13]. Based on these data, the value of the last HbA1C measurement was subsequently identified among the measurements made in the clinical history.

In the subsequent phase, patients were placed to a designated area within the hospital to conduct the ABI measurement. The limb with the lowest ABI value, indicating a more severe vascular compromise, was chosen based on the selection criteria.

For this calculation, the patient was evaluated in the supine position, measuring the systolic pressure on the projection of the brachial artery at the level of the elbow flexure of both upper limbs. Then the systolic pressures were measured in the projection of both lower limbs’ pedal and posterior tibial artery.

Based on these values, the ABI was calculated with the quotient of the highest value obtained in the measurement of the posterior tibial and pedal arteries between the highest value of the systolic pressure of the arm.

## STATISTIC ANALYSIS

Quantitative variables were expressed through measures of central tendency and dispersion according to their type of distribution, and qualitative variables were analyzed through absolute and relative frequencies.

For the bivariate analysis, T-student test or ANOVA were used for variables with normal distribution and U-Mann Withney or Kruskal-Wallis otherwise. In the multiple regression analysis, a generalized linear model (GLM), Poisson log, was used to calculate the crude (cPR) and adjusted (aPR) prevalence ratios.

All analyzes were performed with a 95% confidence level and a statistically significant p<0.05 value. The analysis was performed using the statistical program STATA 17.0 licensed 301709002145.

## ETHICAL ASPECTS

The protocol study was approved by the Institutional Research Ethics Committee of Hospital Nacional Guillermo Almenara Yrigoyen - EsSalud, (NIT: 753-2014-1620), as well as by the Institutional Ethics Committee of the Universidad Peruana Cayetano Heredia (UPCH). (Certificate no. 141-28-14). The participants provided written informed consent. No minors where included in this study.

## RESULTS

### Sociodemographic and clinical characteristics of the population

Two hundred twenty-six patients were included. The average age and SD were 67.4 ± 10.5 years. The 64.6% (n=146) were male and 59.3% (n=134) did not currently have a job.

The average HbA1C and SD were 8.7% ± 2.1. As clinical characteristics, the average body mass index (BMI) and SD were 26.8 ± 4.1 kg/m2, the median time with DM was 20 years with an IQR between 13 and 25 years. The 65% (n=147) had arterial hypertension as comorbidity, 74.8% (n=169) dyslipidemia, 20% (n= 45) were obese, 14.6% (n=33) had a history of smoking, and 38.5% (n=87) were sedentary.

Wounds were present in 25.2% of the patients, 71.9% corresponding to moderate grade II according to the San Elian Classification^11^. 58.8% of these wounds presented PAD with an ABI ≤ 0.9. (See Table 1).

**Table 1.**
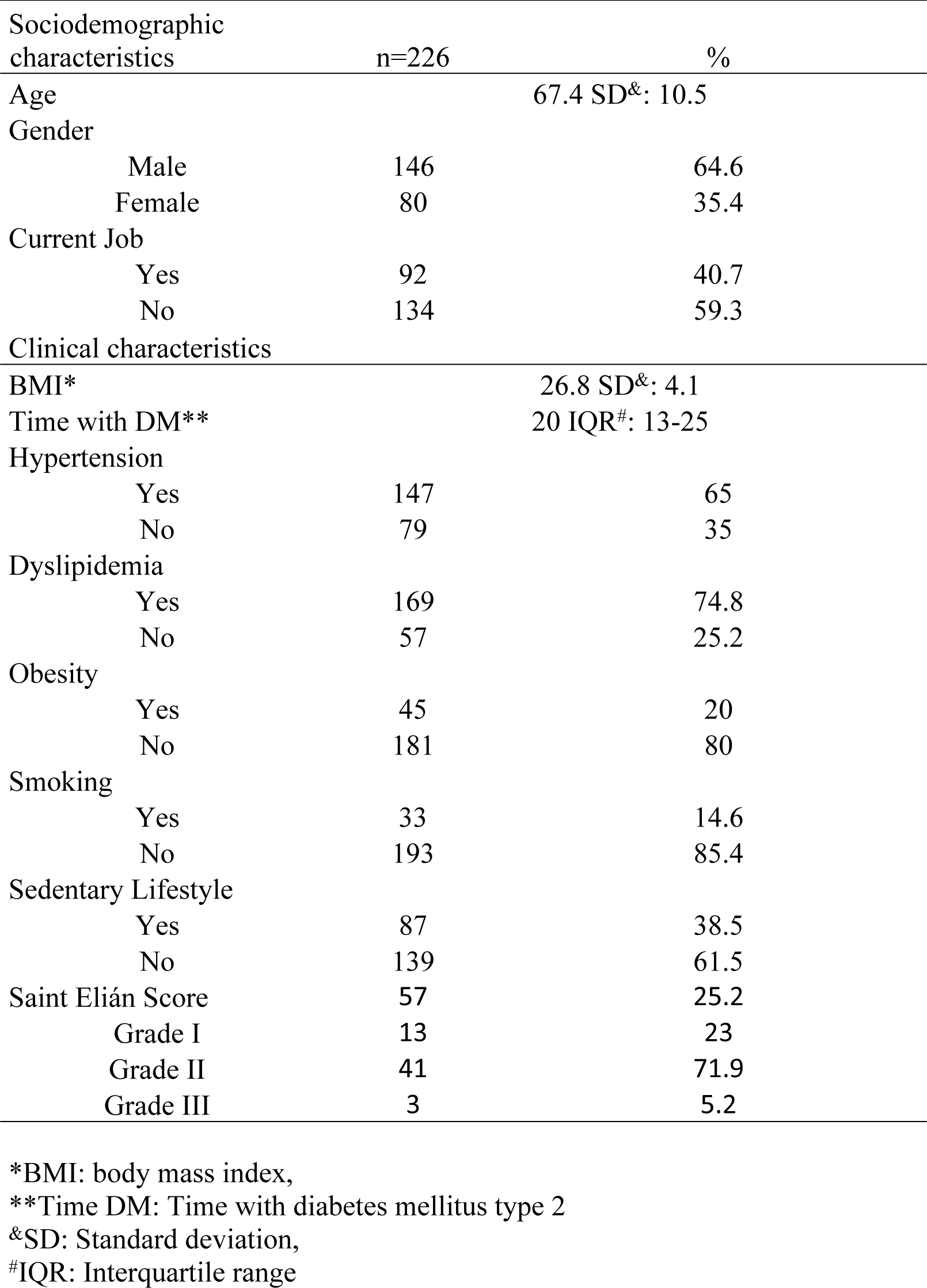
Sociodemographic and clinical characteristics of patients at the HNGAI diabetic foot outpatient clinic during 2015-2017.

| Sociodemographic characteristics | n=226 | % |
| --- | --- | --- |
| Age | 67.4 SD<sup>&</sup>: 10.5 |  |
| Gender |  |  |
| Male | 146 | 64.6 |
| Female | 80 | 35.4 |
| Current Job |  |  |
| Yes | 92 | 40.7 |
| No | 134 | 59.3 |
| Clinical characteristics |  |  |
| BMI* | 26.8 SD<sup>&</sup>: 4.1 |  |
| Time with DM** | 20 IQR<sup>#</sup>: 13-25 |  |
| Hypertension |  |  |
| Yes | 147 | 65 |
| No | 79 | 35 |
| Dyslipidemia |  |  |
| Yes | 169 | 74.8 |
| No | 57 | 25.2 |
| Obesity |  |  |
| Yes | 45 | 20 |
| No | 181 | 80 |
| Smoking |  |  |
| Yes | 33 | 14.6 |
| No | 193 | 85.4 |
| Sedentary Lifestyle |  |  |
| Yes | 87 | 38.5 |
| No | 139 | 61.5 |
| Saint Elián Score | 57 | 25.2 |
| Grade I | 13 | 23 |
| Grade II | 41 | 71.9 |
| Grade III | 3 | 5.2 |

\*BMI: body mass index,
\*\*Time DM: Time with diabetes mellitus type 2
<sup>&</sup>SD: Standard deviation,
<sup>#</sup>IQR: Interquartile range

### Glycemic Control and Ankle Brachial Index

The frequency of good glycemic control was 23% (n=52), and the PAD frequency was 44.7% (n=101, 95%CI: 38.3-51.3). Regarding the latter, 21.8% (n=22, 95%CI: 14.7-31.1) had good glycemic control.

Those who had a normal ABI were 30.1% (n=68, 95%CI: 24.4-36.4) with 30.9% (n=21, 95%CI: 20.9-43.1) with good glycemic control, and 25.2% (n =57, 95%CI: 20-31.3) had an ABI indicative of arterial calcification of the middle layer with 15.8% (n=9, 95%CI: 8.3-28.1) with good glycemic control.

However, no statistically significant association (p=0.126) was found between ABI and glycemic control. (See Table 2).

**Table 2.** Glycemic control and ankle-brachial index of patients at the HNGAI diabetic foot outpatient clinic during 2015 - 2017.

| Glycemic Control | Ankle-Brachial Index (Yao) |  |  |  |  |  |  | P-value* |
| --- | --- | --- | --- | --- | --- | --- | --- | --- |
|  | Total | PAD ( ≤ 0.9 ) |  | Normal (0.91 - 1.3) |  | Calcified (> 1.3) |  |  |
|  | n (%) | n (%) | 95% CI | n (%) | 95% CI | n (%) | 95% CI |  |
| Yes (< 7%) | 52 (23.0) | 22 (21.8) | 14.7 - 31.1 | 21 (30.9) | 20.9 - 43.1 | 9 (15.8) | 8.3 - 28.1 | 0.126 |
| No (≥ 7%) | 174 (77.0) | 79 (78.2) | 68.9 - 85.3 | 47 (69.1) | 56.9 - 79.1 | 48 (84.2) | 71.9 - 91.7 |  |
| Total | 226 (100.0) | 101 (44.7) | 38.3 - 51.3 | 68 (30.1) | 24.4 - 36.4 | 57 (25.2) | 20 - 31.3 |  |

### Relationship between glycemic control and peripheral arterial disease

A generalized linear model was used to calculate the Prevalence Ratios (PR). For the bivariate and multivariate analysis, patients who reported an ABI > 1.3, who were considered “calcified”, were excluded.

In the crude model, HbA1C and PAD were not found to be associated. An association was found between the variables age and sedentary lifestyle with PAD. The non-adjusted model between gender, obesity, arterial hypertension, dyslipidemia, and smoking with PAD did not prove to be statistically significant.

The adjusted model found statistically significant associations for glycemic control, age, and sedentary lifestyle. Regarding age, it was found that for each additional year of life, the probability of having PAD increases by 2% (aPR: 1.02, 95% CI: 1.01-1.03, p=0.005).

And similarly in the case of sedentary lifestyle, those sedentary lifestyle patients had 1.4 times the probability of having PAD compared to those who were not sedentary (aPR: 1.4, 95% CI: 1.1-1.78, p=0.007).

Finally, after performing the statistical adjustment with covariates such as age, gender, obesity, arterial hypertension, dyslipidemia, smoking, and sedentary lifestyle, it was found that HbA1C remained independently associated with PAD, finding that those patients with poor glycemic control had 34% higher probability of presenting PAD compared to those who had good glycemic control (aPR: 1.34, 95%CI: 1.01-1.79, p=0.045). (See Table 3).

**Table 3.**
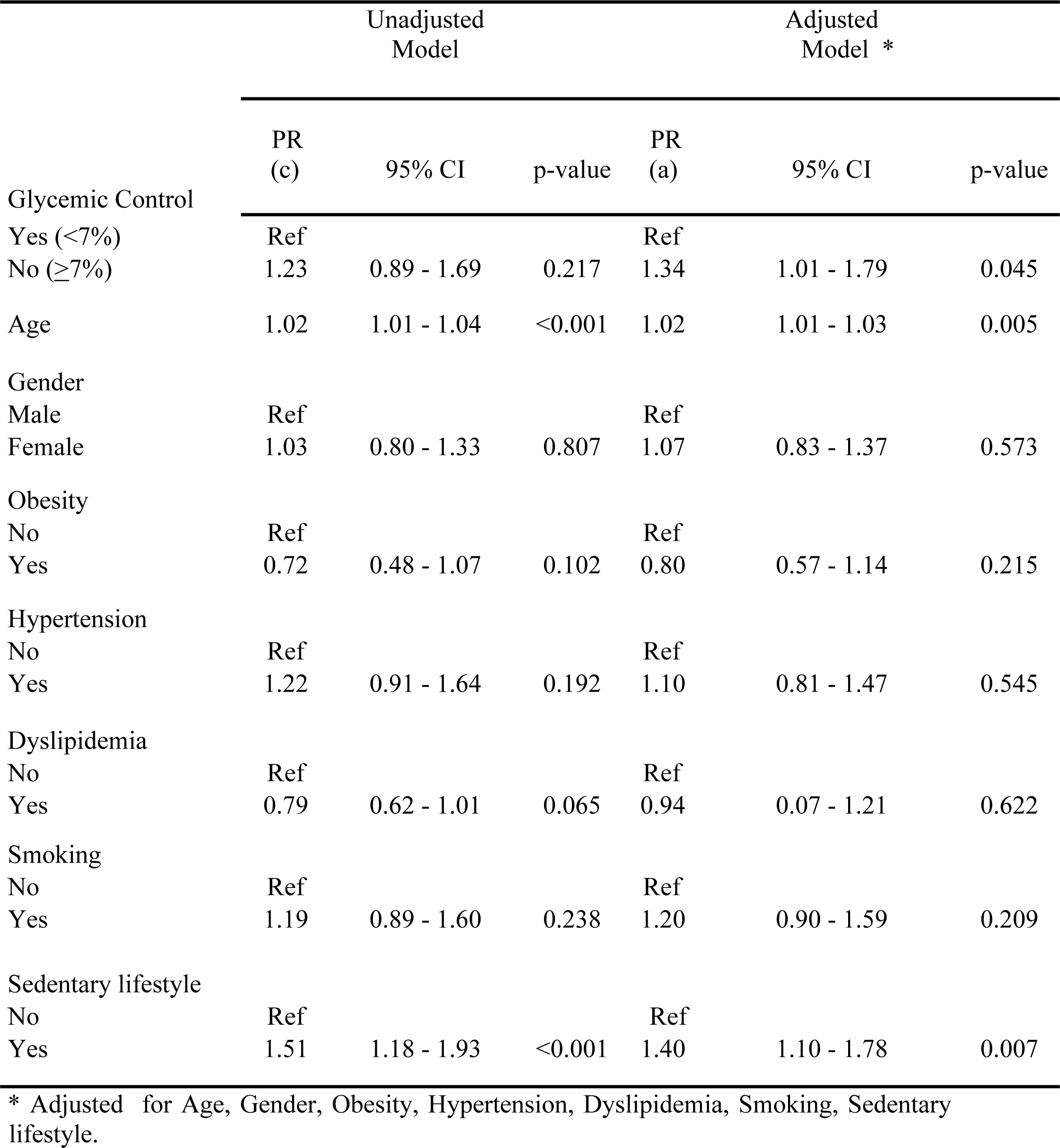
Generalized linear model to evaluate the relationship between glycemic control and PAD of patients at the HNGAI diabetic foot outpatient clinic during 2015-2017.

## DISCUSSION

DF is a macrovascular complication of DM that constitutes a public health problem due to its frequency, mortality, disability, high costs, and high rate of amputations. The study reported 23% good glycemic control from the last HbA1C evaluation, which is similar to findings in studies conducted by Kim et al (27.1%)[14], and Brož et al (33.4%)[15].

In the case of Peruvian literature, the results are similar with 31.8%[16] in patients attended by outpatient clinics and 27%[3] through hospital epidemiological surveillance. The conditions that may contribute to this low prevalence could be due to the lack of adherence of the patients to the treatment, the lifestyle, the type of treatment used, the resistance of the patients to achieve lower HbA1C levels, the different technical aspects in its measurement among other factors that should be explored.

In relation to the Ankle-Brachial Index (ABI), we observed a PAD prevalence of 44.7% (n=101, 95% CI: 38.3-51.3). This prevalence varies when compared to other populations. For instance, studies by Charles et al reported a prevalence of 9.1%[17], Shukla et al found 36%[18], and a study in Peru by Ayala et al documented a notably higher rate of 78%[19]. Furthermore, among those with poor glycemic control, 78.2% (n=79, 95%CI: 68.9-85.3) were found to have PAD. However, the bivariate analysis did not indicate a significant association between ABI and glycemic control.

In this study, the prevalence of arterial calcification, as determined by the ABI, was notably higher in the group with poor glycemic control (27.6%) compared to those with good glycemic control (17.3%). These arterial calcifications, commonly called Monckeberg calcifications, characterize the calcification of the arterial media layer. This process can lead to reduced perfusion to the lower extremities, and its prevalence in certain studies has been reported to range between 58% and 84%[20]. However, it’s important to note that despite our findings, further diagnostic tests like the toe-brachial index or plethysmography are necessary to determine the presence of PAD conclusively[13,20].

In our study, an association was found between age and the probability of having PAD, which is consistent with studies conducted by Kuo-Chin et al[21] (OR 1.04, 95%CI: 1.00-1.07, p<0.05), and that of Zhang et al[22] (OR 1.56, 95%CI: 1.21-1.89, p=0.006) that show that age is an independent risk factor for PAD, as well as that carried out by Martínez-De Jesús et al[13] where age was associated with a higher average ischemia (p<0.01).

We found an association between a sedentary lifestyle and the probability of having PAD, similar to what was reported in the study carried out by Unkart et al[23] where sedentary time had a significant general association with PAD (p=0.048), and this is supported by the fact that prolonged exposure to sitting may play an important role in the development of atherosclerosis in the lower extremities.

In the case of glycemic control, those patients with poor glycemic control had 34% higher probability of present PAD than those with good glycemic control (aPR: 1.34, 95%CI: 1.01-1.79, p=0.045).

There are studies that report the existence of a significant association between HbA1C and PAD in diabetics, such as the one carried out by Muntner et al [24] (OR 2.74, 95%CI: 1.25-6.02, p=0.008), Selvin et al[25] ( RR 4.56 95%CI: 1.86-11.18, p < 0.001), Seong-Woo et al [26] (OR 3.75, 95%CI: 1.30-10.81), Ishii et al[27] (HR 1.63, 95%CI: 1.17-2.28, p=0.0038), Solanki et al[28] (OR:3.00, 95%CI:1.09-8.24, p=0.033), a study conducted by Xiang et al[29] reported a 78.5% prevalence of PAD in patients with DF ulcer in the group with poor glycemic control, compared with a 19.8% in the good control group.

DM is an important risk factor for PAD because hyperglycemia has a major role in the phenomenon of non-enzymatic glycation of proteins and the formation of advanced glycation end-products, leading to vascular inflammation and endothelial cell dysfunction, thereby which promotes greater susceptibility of the vascular bed to atherosclerosis, and the development of PAD in the lower extremities[30].

## CONCLUSIONS

In our study, a minority DF patients exhibited good glycemic control (23%). Moreover, a considerable subset of extremities showed evidence of PAD (44.7%). The individuals with poor glycemic control (HBA1C ≥ 7%) were 34% more predisposed to PAD than those with good glycemic control. Our results emphasize the robust independent relationship between glycemic control and the presence of PAD in DF patients within this population. These findings accentuate the importance of rigorous glycemic control in mitigating PAD associated risks in DF patients.

## LIMITATIONS

By performing a non-probabilistic sampling for convenience as well as the cross-sectional design, it does not allow to establish inferences about causality or to establish the real temporal relationship between glycemic control and the presence of PAD.

HbA1C is a reliable biomarker to monitor glycemic control in diabetic patients, it reflects the accumulated glycemic history of the previous 2 to 3 months, therefore it does not constitute the historical value for each patient. Glycemic control, assessed by HbA1C, is measured on a single occasion, without taking into account its variability over time, so it could be biased. The nature of the study demands prudence in any attempt to extrapolate the results.

## Data Availability

Data file is available from this link: https://docs.google.com/spreadsheets/d/1O4n8q7s3OYyh2yjXQcVvK79HNEEErPYE/edit?usp=sharing&ouid=116904171629274874313&rtpof=true&sd=true

## FINANCING

The study has been self-funded.

## ACKNOWLEDGMENTS

To Dr. Segundo Seclén Santisteban for his constant encouragement.

To Dr. Germán Valenzuela Rodriguez and Dr. Franco Mío Palacios for their support in bibliographic searches.

